# Prognostic Impact of Human Papillomavirus Infection on Cervical Dysplasia, Cancer, and Patient Survival in Saudi Arabia: A 10-Year Retrospective Analysis

**DOI:** 10.1101/2020.05.22.20110247

**Authors:** F.S. Alhamlan, D.A. Obeid, H.H. Khayat, A.M. Tulbah, I.A. Al-Badawi, T.A. Al-Muammar, A.N. Almutairi, S.A. Almatrrouk, M.B. Alfageeh, M.A. Bakhrebah, M.S. Nassar, M.N. Al-Ahdal

**Author notes:** **Correspondence:** F.S. Alhamlan. MBC 03, P.O Box 3354, Riyadh, Saudi Arabia 11211. **Declarations of interest:** none.

## Abstract

Cervical cancer is caused by persistent human papillomavirus (HPV) infection. However, HPV prevalence data and survival rates among HPV-infected women are scare in Saudi Arabia. This study assessed the prevalence of HPV genotypes in a 10 year time-frame. Cervical biopsy specimens underwent HPV detection, HPV viral load using qPCR, HPV genotyping, p16^INK4a^ expression measurement using immunohistochemistry. Kaplan-Meier plots were constructed to analyze overall survival rates. Of the 316 cervical specimens examined, HPV was detected in 96 (30.4%); 37.3% had cervical cancer; 14.2% cervical intraepithelial neoplasia (CIN) III, 4.1% CIN II, and 17.0% CIN I. A significant association was found between HPV-16 viral load and disease progression (*P* < .001, Mann-Whitney *U*) and between HPV presence and cervical cancer (χ^2^, 56.78; *P* < .001). The expression of p16^INK4a^ was a significant predictor of survival: women who had p16^INK4a^ overexpression had poorer survival rates (multivariate Cox regression, hazard ratio, 3.2; 95% CI, 1.1-8.8). In addition, multivariate models with HPV status and cervical cancer diagnosis showed that HPV status was a significant predictor of survival: HPV-positive women had better survival rates than HPV-negative women (haza. These findings suggest that implementing cervical cancer and HPV screening programs may decrease cervical cancer rates and improve survival rates of women in Saudi Arabia.

## 1 Introduction

Human papillomavirus, of the family *Papillomaviridae*, is present worldwide and is known to cause cervical cancer. More than 180 HPV genotypes have been identified, with low-risk types associated with benign hyperproliferative lesions or genital warts and high-risk types associated with premalignant and malignant cervical lesions [1]. Of the 190 million women globally who received a diagnosis of an HPV clinical infection in 2012, approximately 528,000 also received a new diagnosis of cervical carcinoma, and 266,000 women died of this cancer [2]. Approximately 85% of detected HPV cases occur in less developed countries despite current efforts to provide information about this virus and its relationship to cervical cancer to the general public as well as to health care professionals in these countries. Although it is has been estimated that 2.2% of the women in Western Asia have a cervical HPV infection, observations on the HPV burden in the more limited region of Saudi Arabia remain controversial [3]. The results of some studies indicate that the rate of HPV infection in Saudi Arabia is the lowest in the world, whereas other studies show that the rate is high [4], [5], [6], [7].

Every abnormal or dysplastic lesion of the cervix should be considered potentially malignant and may develop into cervical cancer [8], [9]. Initial abnormal cervical epithelial cell detection is generally conducted through a Papanicolaou (Pap) test, commonly known as a Pap smear, which is considered the “gold standard” test, with the results reported using the Bethesda System of classification (Apgar et al, 2003). A biopsy is performed, and histologic testing is used to confirm a diagnosis, with results, listed in order from the mildest to the most severe disease progression, of cervical intraepithelial neoplasia (CIN) I, CIN II, CIN III, and cervical cancer [10], [11]. However, the interpretation of these results is subject to intraobserver and interobserver variability [12]. This subjectivity can be ameliorated by examining the expression of biomarkers in biopsied specimens to identify women with ambiguous results who should be referred for colposcopy and treatment, to distinguish transforming from productive HPV infections, and to predict disease severity [13]. One such biomarker is p16INK4a, a cyclin-dependent kinase inhibitor that coordinates the cell cycle (G1 to S phase) and acts as a tumor suppressor. Overexpression of p16^INK4a^ protein has been used as a biomarker to distinguish transforming HPV infections and, in conjunction with histologic assessment of cervical biopsies, to confirm ambiguous results [14].

Because of the high prevalence of HPV, persistence of HPV infections, and long latency for the development of cervical cancer, primary effective screening for detection of HPV infection and for HPV genotyping during the same clinical visit is important. Such screening would provide better outcomes for all women but especially for those in developing countries. Testing would be for the presence of HPV or HPV-associated serologic markers (HPV biomarkers) in addition to liquid-based cytology, the visual inspection of the cervix, or histologic examination [12], [15].

Cervical cancer is the most preventable cancer, and women with cervical cancer have a high survival rate when it is detected in the early stages [16]. However, although the number of cases of cervical cancer is low in Saudi Arabia, most Saudi women do not seek medical attention until the late stages of the disease. Alhamlan et al. (2017) reported that nearly half of all women with cervical cancer in the Gulf Cooperation Council countries, which include Saudi Arabia, presented at the hospital with regional or distant metastasis stages [16]. They found that only 28% of patients in these countries sought medical care over a 14-year period for early-stage cervical cancer, whereas more than 50% presented to the hospital in late stages, decreasing their survival rate. Specimen were collected from King Faisal Specialist Hospital and Research Centre (KFSHRC), which is a tertiary care and referral hospital in Saudi Arabia. In Saudi Arabia, population cervical screening programs are not available, and no HPV vaccination programs are implemented yet. An overall goal for the present study was to highlight the importance of HPV infection burden and cervical cancer in Saudi Arabia, should our results show this to be the case.

The present study determined the prevalence of HPV and its genotypes among women attending a Saudi Arabian referral hospital between 2006 and 2016, assessed the association of HPV genotypes with cervical dysplasia and cancer, and determined whether HPV presence predicted survival.

## 2 Materials and Methods

### 2.1 Cervical specimen collection

Biopsied formalin-fixed paraffin-embedded (FFPE) cervical specimens at various stages of cervical dysplasia and cancer archived from 2006 to 2016 were obtained from the College of American Pathologists–accredited Pathology Department of KFSHRC, the largest central and referral hospital in Saudi Arabia. All FFPE specimens with abnormal cervical findings were eligible for inclusion, but those with no clinical or demographic data available or those without diagnostic confirmation were excluded. In total, 316 specimens were included in this study.

### 2.2 Cervical specimen processing

The FFPE specimen blocks were processed using a standard method. Briefly, four sections of tissues (4 μm thick) were obtained from each block for molecular assays. To minimize contamination, the microtome blade was disinfected after each use, and an empty paraffin-embedded block was used immediately after each disinfection step to control for any carryover contamination or inhibitors.

Immunohistochemical assays were performed to detect levels of p16^INK4a^ expression in the cervical specimens using an anti-p16^INK4a^ antibody (sc-460, Santa Cruz Biotechnology, Dallas, TX, USA) by following a previously published protocol [17]. Brown nuclear or cytoplasmic staining was considered positive for p16^INK4a^ expression. The p16^INK4a^ expression level was assigned a value from 0 to 3 as follows: 0, no staining; 1, weak focal staining; 2, strong focal or weak diffuse staining; and 3, strong diffuse staining [17], [18].

DNA extraction was conducted using a QIAamp DNA FFPE Tissue kit and following the manufacturer’s instructions (Qiagen; Valencia, CA, USA). This kit is specialized to extract and purify DNA from FFPE tissue. The extracted genomic DNA was quantified and checked for purity using a spectrophotometer, where the ratio of the sample absorbance at wavelengths 260 nm and 280 nm is approximately 1.8 (NanoDrop Technologies; Wilmington, DE, USA). To confirm the presence of DNA in each sample, primers for the β-globin gene (i.e., a housekeeping gene) were used.

### 2.3 Demographic and clinical statistical analyses

Demographic and clinical data were collected, including patient age, marital status, religion, nationality, and specimen pathologic results. All data were collected and analyzed using SAS (version 9.4) software. The percentage of women in each category who were HPV-positive was calculated and is presented with corresponding 95% confidence intervals (CIs). Descriptive analyses were performed for HPV prevalence, HPV genotypes, patient age distribution, potential risk factors, and HPV status. The *χ*^2^ test of independence was used to assess the association between categorical variables. Mann-Whitney and *t* tests were used to assess differences between categorical groups (i.e., HPV status, p16^INK4a^ expression level, and cervical cancer status) and numerical variables, such as age and viral load. Odds ratio estimates and their 95% CIs were generated using the Mantel-Haenszel method. The type I error rate was set at 5%. An exploratory analysis was performed to assess the association between HPV status and age, and the adjusted odds ratio (adjusted for factors that are associated with the risk of HPV infection) was calculated using a multivariate logistic regression model.

The collected data were also used to calculate a survival rate in this hospital-based cohort study. The overall survival rate was defined as the time between the date of diagnosis and the date of death or of the last follow-up observation. The Kaplan-Meier method was used to generate HPV-specific survival curves, and the differences between groups (HPV-positive vs. HPV-negative) were analyzed using the log-rank test. A multivariate Cox proportional hazards analysis was used to evaluate the association of overall survival with each prognostic factor (e.g., HPV type, disease stage, and age). In this cohort analysis, 29 patients overall died within the study time frame. Thus, owing to the small sample size, these results should not be generalized to the general population.

### 2.4 HPV polymerase chain reaction detection

Well-established sets of polymerase chain reaction (PCR) primers were utilized for HPV detection. The primer sets used were MY 09/MY 11 and GP5+/GP6+ which target sequences located within the L1 region of HPV genome [19]. Primer sequences for the first round are as follows MY09 (5’ CGT CCA AAA GGA AAC TGA GC 3’), MY11 (5’ GCA CAG GGACAT AAC AAT GG 3’) with an amplicon size of 450 bp. The primer sequences for the nest PCR are as follows: GP5+ (5’ TTT GTT ACT GTG GTA GAT ACT AC 3’) and GP6+ (5’ GAA AAA TAA ACT GTA AAA CAT ATT C 3’) with an amplicon size of 150 bp. Positive controls (i.e., HeLa cells for HPV-18 and SiHa cells for HPV-16), a negative control (UltraPure DNase/RNase-free water), and internal control primers for the β-globin gene (Takara, Cat. #3868, GH20-forward and PC04-reverse; amplicon size 248 bp) were used. For the first round of PCR, a total reaction volume of 25 uL was prepared that included 1 uL of the template DNA, MY09/11 primers (1 uL each), 12.5 uL of GoTaq Green Master Mix (Promega; Madison, WI, USA), and 9.5 uL of DNase/RNase-free water. For nested PCR, 2 uL of the product from the first round was amplified with GP5/GP6 primers (1 uL each),12.5 uL of GoTaq Green Master Mix (Promega), and 8.5 uL of DNase/RNase-free water. The products from the nested PCR were separated using 1.5% agarose gel electrophoresis and visualized using ultraviolet light (Gel Doc EZ Gel; Bio-Rad).

### 2.5 HPV detection and genotyping using the GenoFlow HPV array test

The GenoFlow HPV array test (DiagCor Bioscience Inc., Hong Kong) is a reverse dot blot assay with the ability to genotype 33 types of HPV. This includes 17 high-risk HPV genotypes (16, 18, 31, 33, 35, 39, 45, 51, 52, 53, 56, 58, 59, 66, 68, 73, and 82), and 16 low-risk HPV genotypes (6, 11, 26, 40, 42, 43, 44, 54, 55, 57, 61, 70, 71, 72, 81, and 84), and a universal probe for the detection of 33 HPV genotypes and out-of-panel HPV genotypes. The test uses a modified PGMY primer set to amplify the L1 region of HPV. A rapid flow-through process enables the hybridization of the PCR products to the 33 HPV genotyping probe spots, a universal probe spot for detecting HPV genotypes outside the panel, or some HPV variants adhered to a membrane. An internal control (the albumin gene) as well as a negative control (UltraPure DNase/RNase-free water) was used along with the samples. The GenoFlow array testing used for genotyping was performed on only the PCR-positive samples because overall cost was a consideration for this study. The genotyping results were interpreted using the manufacturer’s instructions as follows: a valid HPV-positive result was one that included visible signals at the universal, hybridization control, and amplification control probe spots; a valid HPV-negative result included signals at the hybridization control and amplification control probe spots.

### 2.6 Viral load assay

To quantify HPV-16 and HPV-18 DNA levels, multiplex quantitative real-time PCR was performed using a 7500 Fast Real-Time PCR system and software (Applied Biosystems, California, USA). Viral load assays for HPV-16 and HPV-18 were conducted for all of the HPV-positive cases, regardless of the GenoFlow results. The quantitation plasmid used contained the three target sequences of interest: HPV-16, HPV-18, and glyceraldehyde-3-phosphate dehydrogenase (GAPDH) genes. Primers and probes targeted the L1 gene region and were custom designed by Applied Biosystems (Warrington, UK; kit assay #AIY90LA for HPV-18 and #AIX02E2 for HPV-16). Genomic DNA and HPV plasmids were amplified using TaqMan Universal PCR Master Mix (Applied Biosystems; Warrington, UK). The amplification conditions for GAPDH, HPV-16, and HPV-18 were 50°C for 2 min and 95°C for 12 min followed by 50 cycles of 95°C for 15 s and 55°C for 30 s. Positive controls were a SiHa cervical carcinoma cell line that was positive for HPV-16 and a HeLa cell line positive for HPV-18. The negative control was UltraPure DNase/RNase-free water. Tenfold plasmid dilutions were used to construct 5-point standard curves for all three target sequences, and the viral load was normalized to the input amount of cellular DNA. All samples were analyzed three times, and the mean values were expressed as the mean of HPV copies per microliter.

### 2.7 Ethical approval and participant informed consent

This study was conducted according to the World Medical Association Declaration of Helsinki. The study protocol was approved by the Research Advisory Council (Ethics Committee) at KFSHRC (No. 2150001). The need for obtaining written informed patient consent was waived by the ethics committee for the present study because the cervical biopsies were archived, and the collected patient data were coded and thus deidentified.

## 3 Results

The demographic and clinical characteristics of the 316 women included in this study are shown in Table 1. The women were aged 11 to 95 years (mean, 49.5 years; standard deviation, 13.4 years). The prevalence of HPV increased with age from 11-30 years to 46-60 years, with the prevalence for women older than 60 years less than that for women aged 31-45 years. Most of the women claimed Saudi nationality, while 13.6% were non-Saudi. In addition, 74.4% of the women were married, 11.7% were widowed, 10.1% were single, and 3.8% were divorced. The pathologic results indicated that 37.3% of the collected samples were positive for cervical cancer, 14.2% were classified as CIN III, 5.1% as CIN II, and 17.1% as CIN I, and 26.3% were considered normal tissue. HPV status was analyzed with the other demographic and clinical data (age group, religion, nationality, marital status, histology grades, and cervical cancer diagnosis) using the *χ*^2^ test. Marital status, age groups, histologic grades, and cancer diagnosis were significantly associated with HPV status (*P* < .05 for each).

**Table 1.** Demographic and clinical data analyzed by HPV status and presented as percentages of each subcategory (N = 316).

| Category | HPV-positive (n =<br>96)<br>No. (%) | HPV-negative<br>(n = 220)<br>No. (%) | Chi-square (*P* value) |
| --- | --- | --- | --- |
| **Age, years** |  |  |  |
| 11-30 | 2 (11.8) | 15 (88.2) | 8.3 (0.04) |
| 31-45 | 30 (26.3) | 84 (73.7) |  |
| 46-60 | 37 (30.3) | 85 (69.7) |  |
| >60 | 27 (42.9) | 36 (57.1) |  |
| **Mean (SD)** | 52.51 (13.4) | 48.31 (13.4) | *t* test = 2.5 , *P* value =<br>**.01** |
| **Religion** |  |  |  |
| Muslim | 90 (31.8) | 193 (68.2) | 2.6 (.11) |
| Non-Muslim | 6 (18.2) | 27 (81.8) |  |
| **Nationality** |  |  |  |
| Saudi | 87(31.9) | 186 (68.1) | 2.1 (.15) |
| Non-Saudi | 9 (20.9) | 34(79.1) |  |
| **Marital status** |  |  |  |
| Married | 66(28.1) | 169 (71.9) | 9.4 (.02) |
| Divorced | 4(33.3) | 8 (66.7) |  |
| Widowed | 19 (51.4) | 18 (48.6) |  |
| Single | 7 (21.9) | 25 (78.1) |  |
| **Histologic Classification** |  |  |  |
| Normal | 6 (7.2) | 77 (92.8) | 64.5 (<.0001) |
| CIN I | 6 (11.1) | 48 (88.9) |  |
| CIN II | 3 (18.8) | 13 (81.2) |  |
| CIN III | 17 (37.8) | 28 (62.2) |  |
| Cervical cancer | 64 (54.2) | 54 (45.8) |  |
| **Diagnosis** |  |  |  |
| Cancer cases | 64 (54.2) | 54 (45.8) | 50.7 (<.0001) |
| Non-cancer cases | 32 (16.2) | 166 (83.8) |  |

Of the 316 tested cervical specimens, HPV was detected in 96 specimens (30.4%). The most common HPV high-risk types detected were HPV-16 (56.3%), HPV-18 (7.3%), HPV-31 (4.2%), HPV-58 (3.1%), HPV-56 (2.1%), HPV-33 (2.1%), and HPV-45 (1.0%); 19% of the cervical specimens had multiple infections (Table 2). The low-risk HPV types, including HPV-6, 11, 57, and 71, were detected mostly as co-infections with the high-risk types.

**Table 2.** Distribution of high- and low-risk HPV subtypes as detected by GenoFlow HPV array assay in 96 formalin-fixed paraffin-embedded specimens.

| HPV type | No. (%) |
| --- | --- |
| **High-risk Genotypes Detected** |  |
| 16 | 54 (56.3) |
| 18 | 7 (7.3) |
| 31 | 4 (4.2) |
| 33 | 2 (2.1) |
| 35 | 1 (1.0) |
| 45 | 1 (1.0) |
| 56 | 2 (2.1) |
| 58 | 3 (3.1) |
| 82 | 1 (1.0) |
| **Multiple HPV Infections Detected** |  |
| 11, 33 | 2 (2.1) |
| 16, 11, 33 | 1 (1.0) |
| 16, 18 | 3 (3.1) |
| 16, 31 | 1 (1.0) |
| 16, 51 | 1 (1.0) |
| 16, 57, 71 | 2 (2.1) |
| 16, 58 | 1 (1.0) |
| 16, 66, 68 | 1 (1.0) |
| 11, 31 | 1 (1.0) |
| 31, 33, 18 | 1 (1.0) |
| 53, 56 | 1 (1.0) |
| 6, 16 | 1 (1.0) |
| 6, 58 | 1 (1.0) |
| Unknown | 1 (1.0) |
| Negative | 3 (3.1) |

Figure 1 shows the distribution of HPV status (positive or negative) by classification of cervical dysplasia or the presence of cervical cancer. Of the total specimens tested for HPV, 54.2% of cervical cancer cases tested positive, 37.2% of those classified as CIN III tested positive, 18.8% of those classified as CIN II tested positive, and 11.1% of those classified as CIN I tested positive. In addition, 92.8% of the specimens were considered both normal cervical tissue and negative for HPV.

**Figure 1.**
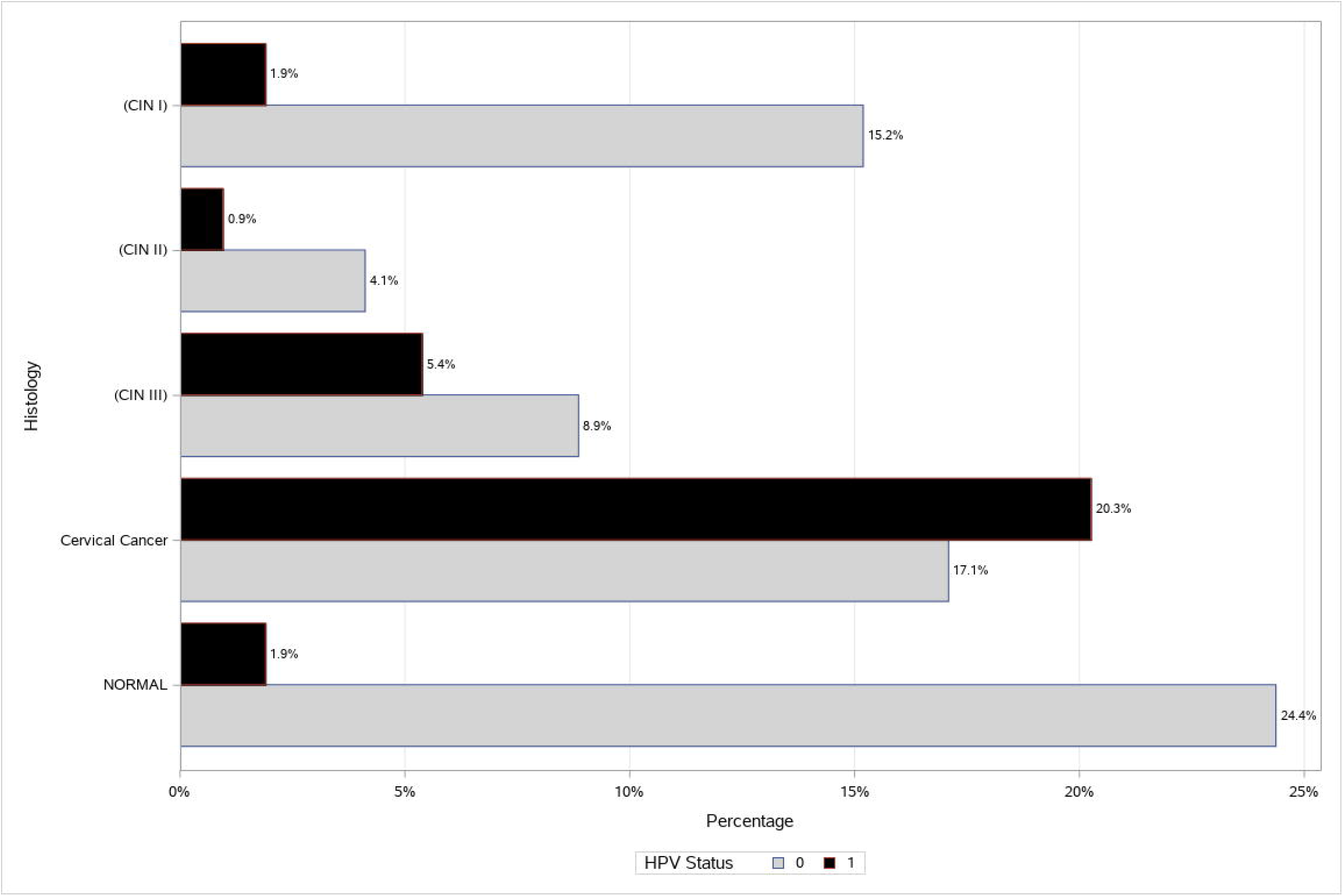
Distribution of the histologic classification by HPV status. HPV status 0 indicates negative for HPV, and 1 indicates positive for HPV. CIN indicates cervical intraepithelial neoplasia. Bars indicate percentages calculated using the grand total number of specimens processed.

The p16^INK4a^ immunohistochemistry staining was strongly positive in 212 samples, and these results are summarized in Table 3. The p16^INK4a^ marker was significantly associated with both histologic abnormalities and HPV status (*P* < 0.0001). For the histologic abnormalities, approximately 86.9% of cervical cancer cases were positive for the p16^INK4a^ marker, and the majority of both CIN II and CIN III specimens were positive for this marker. For the HPV status, 87.5% were both HPV- and p16^INK4a^-positive, and approximately 59.6% were negative for both HPV and p16^INK4a^.

**Table 3.**
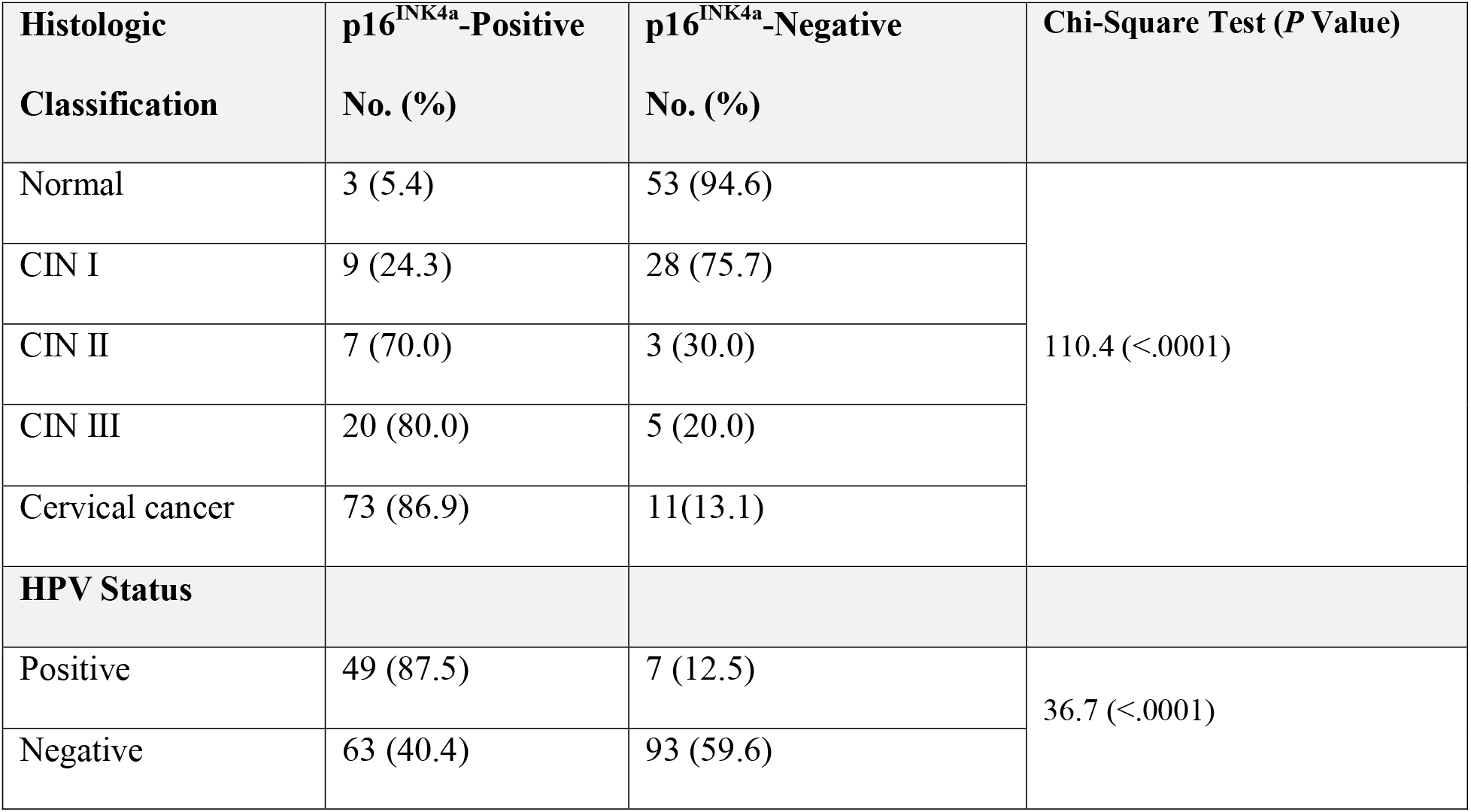
p16^INK4a^ Immunohistochemistry results for 212 samples by histologic classification and HPV status, with percentages calculated using the total number in each subcategory (row).

| **Histologic Classification** | **p16<sup>INK4a</sup>-Positive**<br>**No. (%)** | **p16<sup>INK4a</sup>-Negative**<br>**No. (%)** | **Chi-Square Test (*P* Value)** |
| --- | --- | --- | --- |
| Normal | 3 (5.4) | 53 (94.6) | 110.4 (<.0001) |
| CIN I | 9 (24.3) | 28 (75.7) |  |
| CIN II | 7 (70.0) | 3 (30.0) |  |
| CIN III | 20 (80.0) | 5 (20.0) |  |
| Cervical cancer | 73 (86.9) | 11(13.1) |  |
| **HPV Status** |  |  |  |
| Positive | 49 (87.5) | 7 (12.5) | 36.7 (<.0001) |
| Negative | 63 (40.4) | 93 (59.6) |  |

The survival analysis was conducted using data from 316 women, of which 29 died and 2 were excluded from the analysis. The initial summary of the survival analysis using Kaplan-Meier analysis (Supplementary material, Table S3), indicated that 2 patients had significantly longer survival time, exceeding 5 years, than the remaining patients, who had survival times of 3 years or less; therefore, these two patients were excluded from the analysis. Table 4 summarizes the Kaplan-Meier survival analyses of 314 patients. Figure 2 shows the Kaplan-Meier plot for Cervical cancer clinical groups, Cervical Cancer has a higher risk and lower survival rates than other patients, additional examined groups are shown in the Supplementary Figures S1-S3.

**Table 4.**
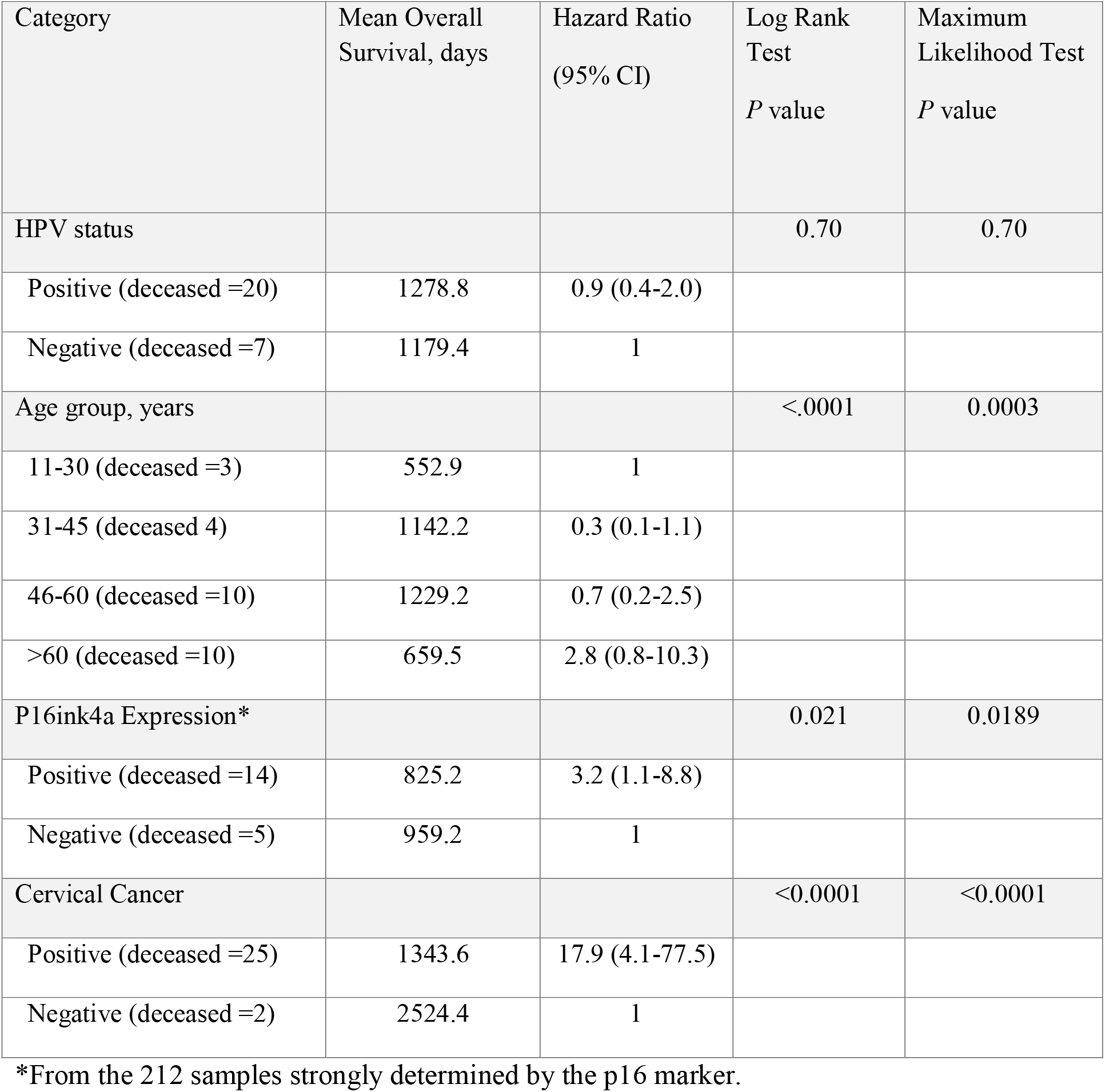
Survival analysis showing Kaplan-Meier analysis and Cox regression analysis summaries.

**Figure 2.**
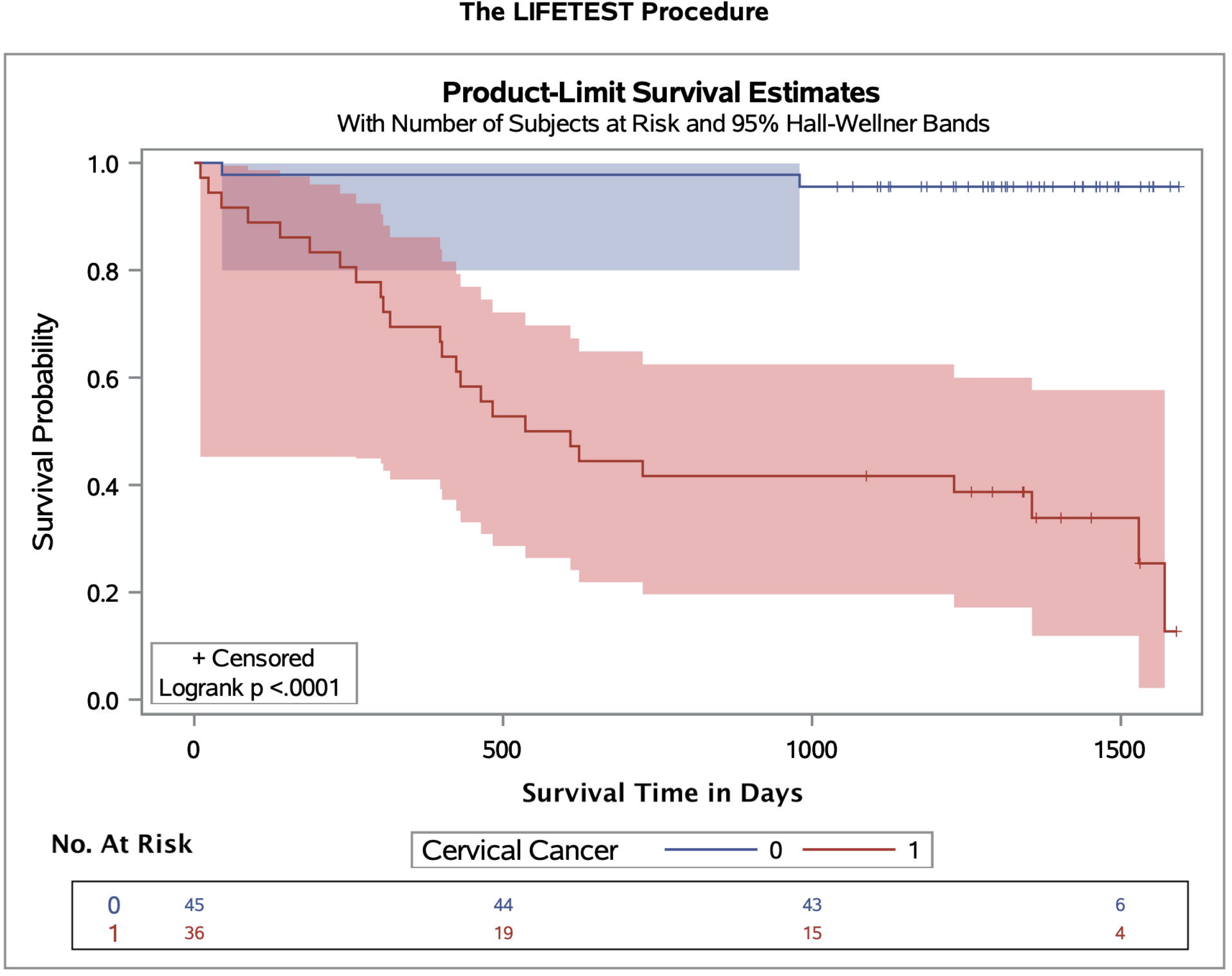
Kaplan-Meier (survival) plot by cervical cancer diagnosis in the KFSHRC cohort, with Hall-Wellner 95% confidence bands and survival rate test. Cervical cancer cases are represented by the red band, and patients negative for cervical cancer are represented by the blue band. The number of the high risk groups are represented below the figure.

The distributions of survival in several groups were tested using the Kaplan-Meier method and validated with the log-rank test (univariate modeling) and using hazard ratios (HRs) validated with the maximum likelihood test. The Kaplan-Meier plots showed that a significant difference in survival distribution was detected by the presence of cervical cancer, age, p16^INK4a^ expression level, and histologic grade (log-rank test, *P* < .05). For the HPV status, a lower HR was detected in positive HPV samples (HR, 0.9; 95% CI, 0.4-2.0). For the age groups, the HR was highest in the oldest group of women (HR, 2.8; 95% CI, 0.8-10.3). For cervical cancer, a higher HR was found in women with cervical cancer (HR, 17.9; 95% CI, 4.1-77.5), and the HR was statistically significant (*χ*^2^, 33.5; *P* < .0001). For the histologic grades, there was insufficient data to conduct an analysis because we did not include patients in the CIN II and CIN III groups who died during the time frame of the study. Regarding the p16^INK4a^ status, a higher HR was found in cervical specimens positive, rather than negative, for p16^INK4a^ expression (HR, 3.2; 95% CI, 1.1-8.8). Kaplan-Meier plots generated for p16^INK4a^ status by survival rate showed that the survival rate was higher among women whose specimens were negative for p16^INK4a^ than among those positive for p16^INK4a^ (log-rank test, *P* < 0.001). This finding indicated that p16^INK4a^ expression directly reflected cervical specimens infected with high-risk HPV and thus added substantial diagnostic accuracy in the evaluation of CIN.

A multivariate Cox regression analysis was conducted to determine the effect of all variables. Using a stepwise method, the model that best predicted survival was one that consisted of HPV status (HR, 0.24; 95% CI, 0.10-0.59) and cervical cancer diagnosis (HR, 39.51; 95% CI, 9.1-171.50). Other models were assessed, and they are described in Table 5.

**Table 5.** Multivariate Cox regression model analyses.

| **A. Multivariate Cox model using cervical cancer and HPV status** |  |  |
| --- | --- | --- |
| **Factor** | **Hazard Ratio (95% CI)** | **P Value** |
| **HPV status** | | $P < .0001$ |
| Positive | 0.2 (0.1-0.6) |  |
| Negative | 1 |  |
| **Cervical cancer** |  |  |
| Positive | 39.5 (9.1-171.5) |  |
| Negative | 1 |  |
| **B. Multivariate Cox model using cervical cancer and p16<sup>INK4a</sup>** |  |  |
| **p16<sup>INK4a</sup> expression** | | $P < .0001$ |
| Positive | 0.5 (0.2-1.6) |  |
| Negative | 1 |  |
| **Cervical cancer** |  |  |
| Positive | 27.9 (5.3-147.1) |  |
| Negative | 1 |  |
| **C. Multivariate Cox model using HPV status, age group, and p16<sup>INK4a</sup>** |  |  |
| **p16<sup>INK4a</sup> expression** | | $P = 0.015$ |
| Positive | 4.3(1.5-12.4) |  |
| Negative | 1 |  |
| **Age group, years** |  |  |
| <30 | 1 |  |
| 31-45 | 1.2 (0.1-11.3) |  |
| 46-60 | 2.3 (0.3-19.5) |  |
| >60 | 12.3 (1.4-106.5) |  |
| **HPV status** |  |  |
| Positive | 0.2 (0.05-0.7) |  |
| Negative | 1 |  |

The distributions of the HPV-16 and HPV-18 viral loads by histologic grade are shown in Figure 3. The viral load assay was conducted on all HPV-positive samples; thus, for HPV-16, the assay was conducted on 70 samples, while for HPV-18, 11 samples were analyzed. The viral load assay detected one more sample than the GenoFlow assay did for HPV-16, whereas for HPV-18, the results were similar to those of the GenoFlow assay. The results of the viral load assay was assessed using the Mann-Whitney test to determine whether there was a significant association between the DNA viral loads for HPV-16 or HPV-18 (measured in DNA copies per microliter) and histologic classification. The distribution of HPV-16 viral load by histologic grade results showed that the highest mean viral load was found in specimens with cervical cancer, followed by those classified as CIN III and CIN II. The distribution of HPV-18 viral load by histologic grade results showed that the highest mean viral load was found in one specimen classified as CIN III, followed by 11 specimens with cervical cancer (Supplementary material Table-S1). There was a statistically significant association of HPV-16 viral load with cervical cancer. The median HPV-16 viral load in the cervical cancer specimens was 5180.6 copies/μl (n = 49), whereas the median of HPV-16 viral load in noncancerous specimens was 331.3 copies/μl (n = 21), indicating a significant association between a high viral load and disease progression (*P* < .001, Mann-Whitney *U*). By contrast, there was no statistically significant association of HPV-18 viral load with cervical cancer (cervical specimens with cancer vs. those without cancer: *P* = .1475, Mann-Whitney *U*). Supplementary material Table S2-A summarizes the HPV-16 and HPV-18 viral loads by cervical cancer status.

**Figure 3.**
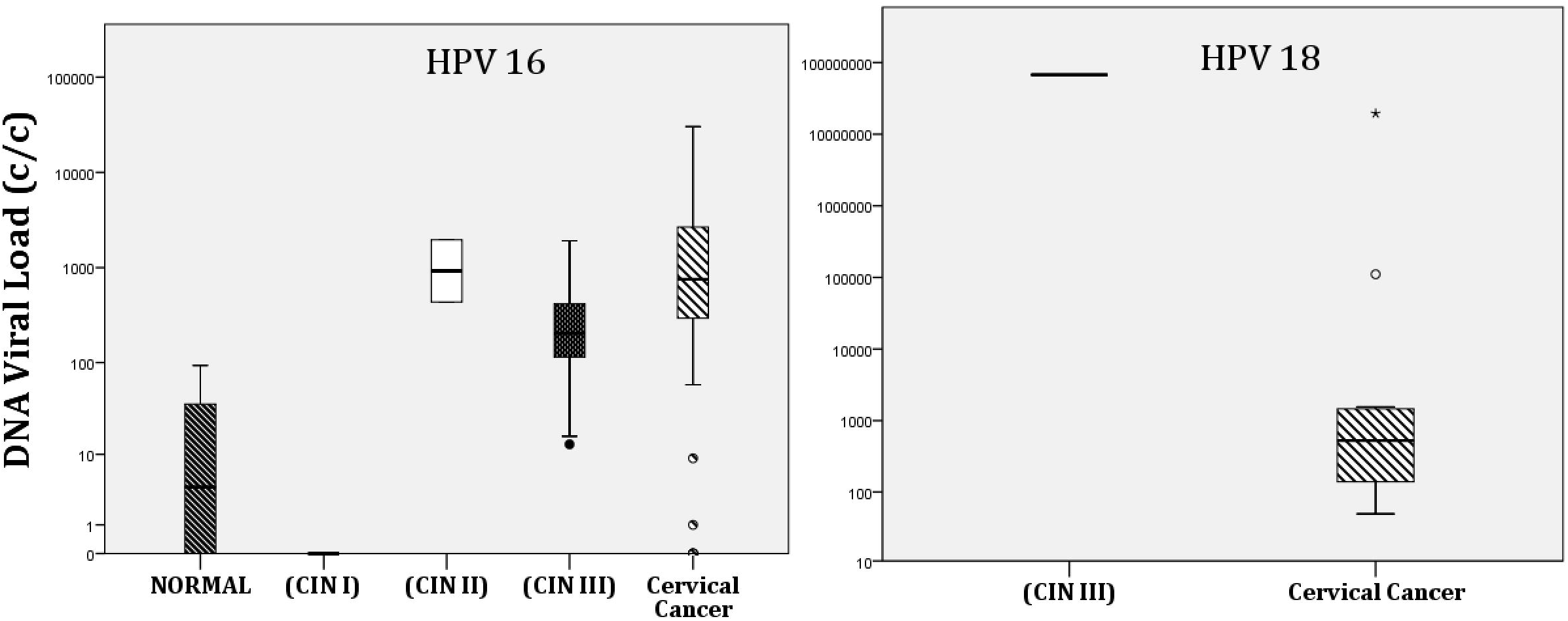
DNA viral loads for HPV-16 and HPV-18 (measured in DNA copies per microliter) distributed by histologic grade. (Left) The highest mean HPV-16 viral load is found in cervical cancer specimens, followed by cervical intraepithelial neoplasia (CIN) III, CIN II, and normal specimens. (Right) The highest mean HPV-18 viral load is found in CIN III specimens and the next highest is in cervical cancer specimens.

A Mann-Whitney test was conducted to determine whether the HPV-16 or HPV-18 viral load was associated with p16^INK4a^ status or a cervical cancer diagnosis. A significant difference was found for the HPV-16 viral load between specimens positive or negative for p16^INK4a^, with positive specimens showing a higher viral load than negative specimens (Supplementary material Table S2-B).

## 4 Discussion

HPV infection is the most common sexually transmitted infection and is particularly common among sexually active young women. The results of the present study showed slight differences in age-specific HPV prevalence, with HPV infection more common in middle-aged women (31-45 years) than in younger women (11-30 years). This discrepancy with the worldwide trend could be attributable to low sample size or differences in sexual practice, lifestyle, and genetic predisposition in Saudi Arabia [16]. However, further studies and evaluation will be required because of the current changes in socioeconomic status, lifestyles, and sexual behaviors in regions that have been traditionally considered to be conservative societies.

HPV was detected in 96 of the 316 cervical specimens examined in the present study. The three most common HPV types detected were HPV-16 (56.3%), HPV-18 (7.3%), and HPV-31 (4.2%), with 19% of the cervical specimens having multiple infections. These data are consistent with previous studies reporting that HPV-16 is the most common type followed by HPV-18 where there was a significant association between the presence of HPV and cervical cancer *χ*^2^, 56.8; df = 1, *P* < .001) [20]. Our study also showed that the GenoFlow assay had less sensitivity than the qPCR assay. In studies in our laboratory, the specificity of the GenoFlow assay is 100%, but its sensitivity is considered low (62%) [21].

The present study also evaluated the association of HPV with the prognosis of women with cervical dysplasia. A multivariate Cox regression analysis showed the model with HPV status and cervical cancer diagnosis to be a significant predictor of survival. In this model, women with HPV-negative cervical specimens had a poorer survival rate than those with HPV-positive specimens. The results of many previous studies agree with our results, and those studies that are not in agreement do not have larger sample sizes than that of the present study. Rodrigues-Carunchio et al. (2105) found that patients with cervical cancer and confirmed HPV negativity had significantly worse disease-free survival than women with HPV-positive tumors. In their multivariate analysis, HPV-negativity and International Federation of Gynecology and Obstetrics staging were associated with increased risk of progression and mortality [22]. Li et al. (2018) published a comprehensive meta-analysis showing the prognosis significance of HPV DNA status in cervical cancer [23]. They evaluated survival data from 2,838 women with cervical cancer and found that women who tested positive for high-risk HPV had a better cervical cancer prognosis (pooled HR, 0.4; 95% CI, 0.3-0.5; *P* < .001) and better overall survival (pooled HR, 0.6; 95% CI, 0.5-0.8; *P* = .001) than those who tested negative for high-risk HPV. Previous studies have discussed the better survival outcomes for women who are HPV-positive and have cervical cancer [24]. Infection with a high-risk HPV is strongly associated with the development of cervical carcinoma. The gene sequences encoding the two major viral oncoproteins, E6 and E7, are consistently retained and expressed in cervical cancer cases. The E6 and E7 genes are thought to play causative roles because E6 promotes the degradation of p53, and E7 binds to the retinoblastoma protein and disrupts its complex formation with E2F transcription factors [24]. However, there are independent factors that cause p53 mutations. The association of p53 mutation with metastases may explain the poor prognosis reported for HPV-negative primary cancers, many of which already contain mutant p53. A high proportion of p53 mutations detected in both primary and metastatic cancers are GC to TA transversions, strongly suggesting a role for external carcinogens in the development of these cancers. The evidence supports that HPV-negative cancers, which frequently carry somatic p53 mutations, show a worse prognosis than HPV-positive, wild-type p53 tumors [25].

Our data showed unexpectedly high numbers of CIN-III and cervical cancer cases that were negative for the presence of HPV. For specimens that had cervical cancer, these results may be explained by cervical cancer tumors that were not induced by the presence of a persistent HPV infection, including the misclassification of endometrial cancers or the metastasis of other cancers to the cervix, the loss of HPV expression, or the existence of cervical cancers that are not induced by HPV [26]. However, many reports have indicated that HPV is a necessary but not sufficient cause of invasive cervical cancer; a variable proportion of tumors have been reported to be negative for high-risk HPV [27]. In addition, a negative HPV test result from a cervical cancer specimen does not necessarily mean that HPV was never involved in the etiology of that cancer; an HPV viral infection may have been involved in the early development of the cancer but thereafter resolved. Nevertheless, we acknowledge that there are limitations in the current testing for detection of HPV, especially between the DNA and mRNA testing approaches. Currently, a DNA-based assay is considered best for HPV detection and typing in early infection stages; however, once the disease progresses, RNA-based assays are considered more accurate [28]. Moreover, to obtain a higher number of samples, we used FFPE specimens, rather than fresh samples, and this may have affected our results. Another limitation was the use of our mentioned DNA detection assay instead of one that uses shorter S-phase-promoting factor primers, which are more suited to FFPE DNA detection. The decision to use the assay we chose was that the availability and the cost of the instruments and reagents were important factors and that previous studies provided PCR and gel confirmations for comparison with our results [17], [18].

The present study evaluated the viral loads in cervical specimens positive for HPV-16 and HPV-18. There was a significant increase in lesion severity with viral load in HPV-16–positive cervical specimens, indicating a significant association between HPV-16 viral load and disease progression. Although the viral load was higher in HPV-18–positive specimens than in the HPV-16–positive specimens, because of the small sample size, HPV-18 viral load was not significantly associated with cervical cancer status. The finding that HPV-16 viral load is positively associated with disease progression is consistent with previous studies. Xi et al. (2011) examined the HPV-16 viral load in a case-control study in which patients were followed up and their HPV viral load quantified [29]. They found that the risk of CIN III increased with increasing HPV-16 DNA load at the follow-up visit. Therefore, the viral load in newly detected infections and changes in the viral load predict persistence and progression of HPV-16 infections. Carcopino et al. (2012) also assessed the clinical significance of HPV-16 and HPV-18 viral loads to determine an optimal prediction of high CIN among patients referred for colposcopy [30]. They found that whereas the median HPV-16 viral load increased significantly with increased lesion grade, HPV-18 viral load showed very poor predictive value. To date, quantification of HPV viral load as a prognostic tool remains controversial because it has often yielded conflicting results. The main reasons for the apparent discrepant results could be the lack of standardized procedures and no standardized expression of viral loads. Therefore, the extent to which HPV viral load quantification can be used in clinical practice must be rigorously assessed in the context of the method used to measure viral load and the type-specific differences observed for cases of CIN II or greater disease progression [31].

The present study also assessed p16^INK4a^ expression in cervical biopsy specimens and found that expression of p16^INK4a^ increased with the severity of histologic abnormality. This finding is consistent with previous studies that reported evidence that p16^INK4a^ immunostaining correlates with the severity of histological abnormalities [32]. Previous studies have shown that 16^INK4a^ is a highly sensitive and specific marker of high-grade squamous and glandular neoplasia of the cervix owing to its overexpression in cancerous and precancerous cervical lesions [33]. However, reproducibility of this finding may be limited by the lack of a standardized method to quantify and interpret p16^INK4a^ immunostaining. Interestingly, the results of the immunohistochemistry biomarker p16^INK4a^ were more strongly correlated with abnormal histologic classification than with HPV status.

To our knowledge, this is the first study examining the association between HPV infection and cervical cancer survival among women in Saudi Arabia. This study used a sensitive method capable of detecting a wide range (33 types) of HPV types, enabling the detection of most HPV types that have been associated with genital and sexual contacts. In addition, the 10-year retrospective analysis is a substantial strength of the study. However, there are several limitations of this study that should be considered when interpreting our results. Because of the low number of cervical cancer cases and late diagnoses, the number of study specimens was relatively small. This small number also informed our decision to use FFPE specimens, rather than fresh samples, to obtain a higher number of samples for the analyses. Although HPV detection assays have their limitations, we took steps to ensure the accuracy of our findings, including using only samples that passed the β-globin amplification internal control test (samples that failed this control test were excluded), running positive and negative controls with the samples, capturing gel images to determine specific amplicon sizes, and evaluating DNA quality using a nanopore sequencing system.

## 5 Conclusions

The present study determined the prevalence of HPV among women attending a main referral hospital in Saudi Arabia between 2006 and 2016. The results suggested that women with HPV-negative cervical specimens may have a poorer mean survival rate than those with HPV-positive specimens. Our study also highlighted the importance of accurate molecular diagnostic techniques for HPV detection and identification, which is crucial for diagnosing patients at risk. These findings contribute to the scientific and medical databases and suggest that implementing cervical cancer and HPV screening programs in developing countries may help control cervical cancer and may improve survival rates.

## Data Availability

Data is available upon request

## Acknowledgements

We thank the Infectious Diseases Program, National Center for Biotechnology in King Abdulaziz City for Science and Technology for their financial and intellectual support. We also thank the members of the pathology department, chaired by Dr. Fouad Aldayel, for their help and efforts to retrieve the archived cervical specimens. We thank Dr. Angela Cox from Sheffield Medical School for her advice on the statistical analysis. The financial support of the Research Centre administration at KFSHRC is highly appreciated.

## Funding/Support

This work was supported by the Infectious Diseases Program, National Center for Biotechnology in King Abdulaziz City for Science and Technology and the Research Centre administration at King Faisal Specialist Hospital and Research Centre.

## Role of the Funders

The funders had no role in study design; in the collection, analysis and interpretation of data; in the writing of the report; and in the decision to submit the article for publication.

## Authors’ contributions

FA is the principal investigator of this study. DO analyzed patient data and survival rates; HK and AA are laboratory technicians who ran the molecular assays. AT, IA, TA are physicians who provided the biopsies and helped in the clinical data interpretation. SA, MF, MB, MN, and MA analyzed and interpreted the patient data regarding the HPV infection and disease progression. All authors helped in writing and reviewing the manuscript. In addition, all authors read and approved the final manuscript.

## Notes

### Competing Interest Statement

The authors have declared no competing interest.

